# Plasma multiomics distinguishes pulmonary tuberculosis from other respiratory infections

**DOI:** 10.64898/2026.03.18.26348671

**Authors:** Zaynab Mousavian, Firdaus Nabeemeeah, Mary M. Nellis, Neel R. Gandhi, Russell R. Kempker, Dean P. Jones, Hadiya Johnson, Mojahidul Islam, Matthew J. Magee, Neil Martinson, Ashish A. Sharma, Jeffrey M. Collins

## Abstract

Novel blood-based biomarkers for tuberculosis (TB) are needed to develop rapid, point-of-care diagnostics. We sought to use combined plasma metabolomics and high-density cytokine profiling to identify a biomarker signature that can differentiate pulmonary TB (PTB) from patients hospitalized with other respiratory diseases and ambulatory household contacts with TB symptoms. We analyzed plasma concentrations of 28 cytokines and 118 metabolites from 391 adults (≥18 years) presenting with respiratory symptoms suggestive of TB, of which 187 had PTB confirmed by Xpert MTB and/or *M. tuberculosis* sputum culture and 204 were controls in whom PTB was excluded. Our study identified a 5-marker signature (IFN.gamma, IL.22, IL.10, methionine and oxoproline) with an AUC of 0.97 (95% CI: 0.95–1.00) in the test set. The signature had 98% and 84% sensitivity at 70% and 98% specificity respectively, which meet WHO target product profiles for both non-sputum triage and diagnostic TB tests.

## Introduction

Tuberculosis (TB) remains a leading infectious cause of death globally, with an estimated 1.23 million deaths in 2024 (1). Early and accurate diagnosis of TB is crucial, as it facilitates timely treatment initiation, which helps limit disease transmission, minimize lung damage and post-TB complications, and reduce mortality (2). While the Xpert MTB-RIF assay demonstrates high sensitivity (∼80%-90%) and specificity (∼98%) in adults (3), implementation in low-resource settings remains challenging due to requirements for good quality sputum samples, laboratory infrastructure, trained staff, electricity, and regular maintenance of machines (4). Moreover, results from a recent meta-analysis estimated that 23% of adults being evaluated for TB cannot produce sputum due to scarcity (5). Limitations of existing standards for TB diagnostics highlight the urgent need for accurate, blood-based diagnostic tools that can be adapted to point-of-care (POC) platforms for use in resource-limited settings.

Advances in high-resolution omics technologies, including genomics, transcriptomics, proteomics, and metabolomics, have accelerated the discovery of blood-based biomarkers for TB diagnosis (6–8). Although blood-based transcriptomic signatures show promise, they require amplification cartridges and laboratory infrastructure, presenting similar barriers to the Xpert MTB/RIF assay. Conversely, antibody-based detection methods used in many POC platforms can often detect protein or metabolite biomarkers (9). Integrating biomarkers across molecular layers, particularly proteins and metabolites, has the potential to improve diagnostic accuracy and robustness by capturing complementary and/or related biological signals. Yet multiomics approaches remain underutilized due to challenges in data integration and the limited availability of comprehensive, multi-modal datasets (6). An additional limitation of TB biomarker studies to date is that control groups are often asymptomatic, which may overestimate diagnostic performance in real world settings. Including controls with non-TB respiratory diseases or symptoms may provide a more clinically relevant and rigorous assessment of biomarker specificity.

In this study, we compared combined plasma metabolomics and cytokine profiles in persons with pulmonary TB (PTB) to a cohort of symptomatic participants hospitalized with a non-TB respiratory illness (10) as well as a cohort of household contacts who reported TB symptoms at the time of evaluation. Our primary objective was to identify and validate combined metabolite and cytokine biomarker signatures for TB diagnosis across care settings that meet or exceed WHO-defined Target Product Profile (TPP) thresholds for non-sputum POC diagnostic tests (>65% sensitivity at 98% specificity) or a TB triage test (>90% sensitivity at 70% specificity) (11). Second, we explored the potential of combined metabolite and cytokine biomarker signatures to predict adverse treatment outcomes. Finally, we examined how metabolic responses interact with cytokine signaling networks.

## Results

### Clinical characteristics of the study participants

We analyzed 391 plasma samples obtained from adults (≥18 years) presenting with respiratory symptoms who were evaluated for PTB using sputum samples tested with Mtb culture and/or Xpert MTB-RIF assay (**Figure 1a**). This cohort comprised 187 individuals in whom PTB was microbiologically confirmed and 204 individuals in whom PTB was excluded (**Table 1**). The PTB group included biobanked plasma samples collected from previous TB clinical trials (n=119) (12, 13) and a South African study of patients hospitalized due to respiratory illness (n=68) (14). The other respiratory disease control group included participants hospitalized due to a non-TB respiratory illness (n=165) and household contacts of people with PTB who reported at least one TB symptom when screened (n=39). Overall, participants ranged in age from 18 to 92 years, and 80 (20%) were people living with HIV (PLWH). Among those with PTB, 41% were female, the median age was 31 years (IQR: 24–41), and 21% were PLWH. In the other respiratory disease control group, 50% were female, the median age was 51 years (IQR: 38–62), and 20% were PLWH. Out of 187 individuals with PTB, 178 had a classified treatment outcome: cure (n = 122) or failure (n = 56) (**Figure 1b**).

**Fig. 1.**
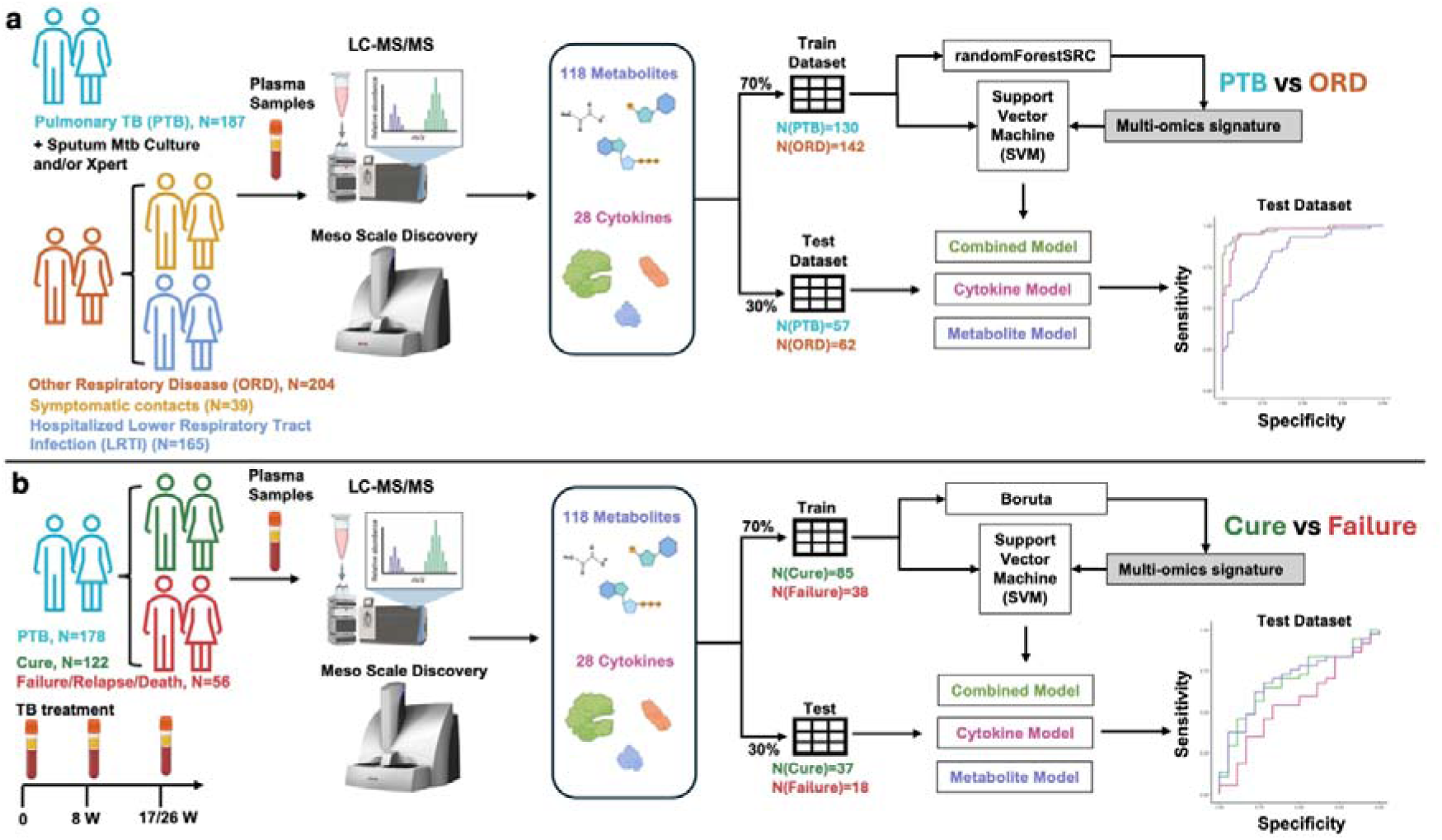
Schematic representation of the study design. Identification of a multiomics signature for **a)** TB diagnosis and **b)** TB treatment response prediction.

**Table 1:** Overview of demographic and clinical characteristics of the study cohort.

| Characteristics | PTB |  |  | ORD |  |  |
| --- | --- | --- | --- | --- | --- | --- |
|  | South Africa |  |  | South Africa |  |  |
|  | Hospitalization Study Samples | TB Clinical Trial Samples | Total | Hospitalization Study Samples | Household contact samples | Total |
| N | 68 | 119 | 187 | 165 | 39 | 204 |
| Age, median (IQR) | 38 (26.75–54.25) | 28 (23–36) | 31(24-41) | 53 (41–62) | 40 (23–64) | 51.5(38-62.25) |
| Female, N (%) | 26 (38.2%) | 50 (42%) | 76(40.6%) | 78 (47.2%) | 24 (61.5%) | 102 (50%) |
| PLWH, N (%) | 29 (42.6%) | 11 (9.2%) | 40 (21.3%) | 32 (19.3%) | 8 (20.5%) | 40 (19.6%) |
| CD4, median (IQR) |  |  | 86(43-286) |  |  | 149(57-469) |
| HIV VL, median (IQR) |  |  | 109401(3358-1175000) |  |  | LDL(LDL-48216) |
| Symptoms, N (%) |  |  |  |  |  |  |
| Cough |  |  |  | 154 (93.3%) | 34 (81%) | 188 (90.8%) |
| Fever |  |  |  | 28 (17%) | 8 (19%) | 36 (17.4%) |
| Loss of weight |  |  |  | 70 (42.4%) | 2 (33.3%) | 72 (42.1%) |
| Night sweats |  |  |  | 42 (25.5%) | 1 (16.7%) | 43 (25.1%) |
| Chest Xray, Cavitation, N (%) | 8(11.7%) | 92(77.3%) | 100(53.4%) | 2 (1.2%) | 0 | 2 (0.99%) |
| Xpert results, Detected, N (%) | 68 (100%) | 79 (66.3%) | 147 (78.6%) | 0 | 0 | 0 |
| Sputum TB Culture results, positive, N (%) | 66 (97%) | 115 (96.6%) | 181 (96.7%) | 0 | 0 | 0 |
| Smear grade |  |  |  |  |  |  |
| 0 |  | 22 | 22 |  | 23 | 23 |
| 1+ |  | 43 | 43 |  |  |  |
| 2+ |  | 52 | 52 |  |  |  |

PTB: Pulmonary TB; ORD: Other respiratory disease; HIV VL: HIV viral load.

### Differentially Abundant Metabolites and Cytokines in PTB vs. other respiratory diseases

We measured plasma concentrations of 118 metabolites and 28 cytokines for all 391 participants. The principal component analysis (PCA) plot demonstrated no batch effect with respect to any variable (**Supplementary Figures 1 and 2**). Each dataset was analyzed using a linear model implemented in the *limma* R package, with sex, age, and HIV status included as covariates. In total, five metabolites were upregulated and 41 were downregulated in the PTB versus the other respiratory disease group (Benjamini-Hochberg (BH) FDR-corrected p-value < 0.05) (**Supplementary Figure 1**). Among these, methionine, oxoproline, and histidine were significantly downregulated in PTB. In the cytokine dataset, 19 cytokines were upregulated and 2 were downregulated in PTB versus other respiratory disease (BH FDR-corrected p-value < 0.05) (**Supplementary Figure 2**). Notably, IFN.gamma and IL.1β were strongly upregulated (log2FC > 2), while IL.10 was downregulated (log2FC < –1) in PTB due to its upregulation in hospitalized pneumonia patients included in the control group.

**Figure 2.**
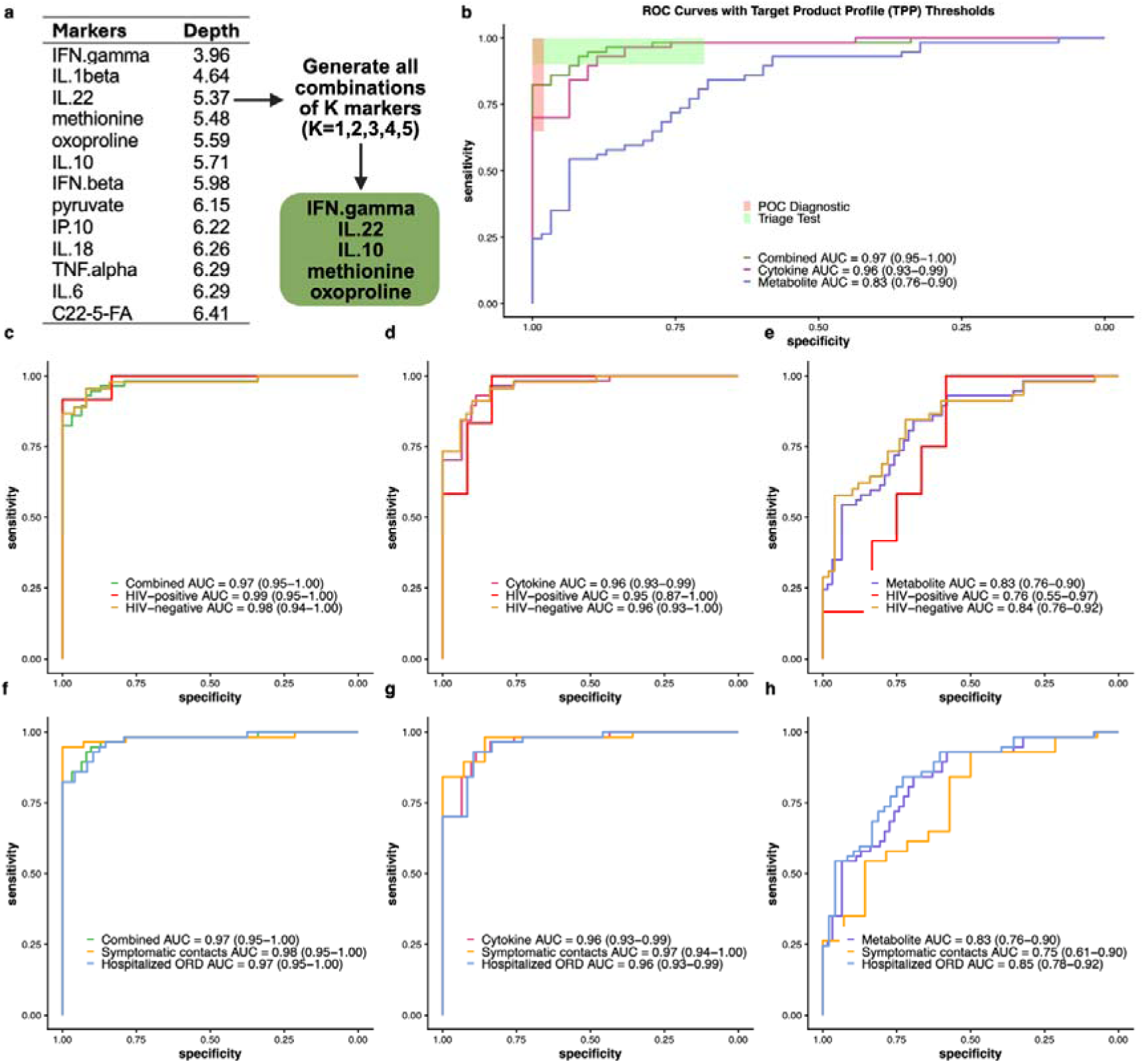
Identification of a multiomics signature for diagnosing PTB versus other respiratory disease. **a)** Selected cytokines and metabolites and their corresponding depths identified by *randomForestSRC*. Depth indicates the relative importance of each variable in classification. **b)** ROC curves for the combined, cytokine, and metabolite models. POC diagnostic and triage test thresholds are indicated by the red and green boxes, respectively. **c-h)** ROC curves for showing the performance of the combined, cytokine, and metabolite models within subgroups stratified by HIV status and by control type (symptomatic contacts and hospitalized ORD). AUC values with 95% confidence intervals are reported for each ROC. ROC: Receiver Operating Characteristic; PTB: Pulmonary TB; AUC: Area Under the Curve; LRTI: Lower Respiratory Tract Infection.

### Identification of a multiomics signature for PTB

We integrated cytokine and metabolite datasets to identify a multiomics signature distinguishing individuals with PTB from other respiratory diseases. For model building, the cohort was split into discovery (70%) and validation (30%) datasets, maintaining the same proportions of PTB and other respiratory disease groups. Specifically, the discovery dataset included 130 PTB and 142 other respiratory disease samples, with 28 HIV-positive individuals in each group (**Figure 1a**). The validation dataset comprised 57 PTB and 62 other respiratory disease samples, with 12 HIV-positive individuals in each group (**Figure 1a**). For biomarker discovery, we applied *RandomForestSRC* (15), a random forest-based feature selection approach, to the discovery dataset. The model identified ten cytokines and three metabolites across different depths, where depth reflects the contribution of each marker to model performance in distinguishing the PTB group from the other respiratory disease group. (**Figure 2a**).

To explore which combination of markers, with a reasonable size for clinical implementation as a biomarker signature, could achieve the best performance, we evaluated all 2,379 possible combinations of Kmarkers (where K = 1,2,3,4,5) from the 13 markers. For ROC analysis, the discovery dataset was used to train support vector machine (SVM) classifier model, which was then tested on the validation dataset. For each combination, we calculated the AUC with a 95% confidence interval, along with the sensitivity at 98% and 70% specificity, each with a 95% confidence interval. Among all combinations listed in **Supplementary Table 1**, a 5-marker signature comprising three cytokines (IFN.gamma, IL.22, and IL.10) and two metabolites (methionine and oxoproline) had high AUC and sensitivity at 98% and 70% specificity when combinations were ranked based on the sum of these three values. Three feature sets were evaluated: (1) the three cytokines, (2) the two metabolites, and (3) the combined five-marker signature. The combined cytokine-metabolite model had an AUC of 0.97 (95% CI: 0.95–1.00) in the test set (**Figure 2b**). Both the cytokine-only and combined model had 98% sensitivity (95% CI: 92%–100%) at 70% specificity. However, at 98% specificity, sensitivity was higher in the combined model at 84% (95%CI: 73%–94%) versus the cytokine-only model [72% sensitivity (95% CI: 58%–89%)] (**Figure 2b**).

Since there was a substantial difference in median age between PTB (35 years) and other respiratory disease (51 years) groups, we evaluated whether the performance of the combined 5-marker signature might be influenced by age or other covariates in addition to TB status. To assess this, we used the SVM classifier score derived from the combined 5-marker signature as the outcome in a linear regression model and included TB group as the primary predictor and age, sex, and HIV status as covariates in the model. The SVM score represents the probability of TB based on the signature score. This approach allowed us to evaluate whether the signature’s classification performance was independent of these potential confounding factors. In this model, the coefficient for the TB group was 0.7 with a highly significant p-value (< 0.001), indicating a strong association between the SVM score and TB status. By contrast, the coefficients for age (−0.001, p=0.2), sex (−0.05, p=0.1), and HIV status (0.05, p=0.1) were small and not statistically significant. These results suggest that the SVM classifier score is driven primarily by TB status and that age, sex, or HIV infection had no meaningful impact on the SVM score.

Next, we evaluated the performance of the three models within subgroups of the validation dataset, stratified by HIV status (HIV-positive vs. HIV-negative) and by type of other respiratory disease controls (hospitalized vs. ambulatory), to assess the robustness of each signature across clinically relevant groups. As shown in **Figure 2c**, the combined model demonstrated consistent performance regardless of HIV status (AUC = 0.99 and 0.98 respectively for persons living with and without HIV), and across both hospitalized and ambulatory participants (AUC=0.97 and 0.98 respectively) (**Figure 2f**). In contrast, both the cytokine- and metabolite-only models showed reduced performance in PLWH (**Figures 2d and 2e**), and the metabolite model also exhibited decreased performance among ambulatory participants (**Figure 2h**).

We also examined the performance of previously published biomarker signatures using exclusively cytokines or metabolites (6, 16, 17). This included a signature using the kynurenine/tryptophan ratio and retinol (17), as well as cytokine-based signatures including IFN-γ, IP-10, TNF-α, and a 3-marker signature consisting of IP-10, IL-6, and IL-10 (6). As shown in Figure 3a, the signature from the present study performed better in distinguishing PTB from other respiratory diseases compared with the kynurenine/tryptophan ratio and retinol (AUC 0.64, 95% CI: 0.54−0.74), though signature performance improved when hospitalized participants were excluded from the control group (Figure 3c), indicating that this signature may only be effective in TB screening or outpatient care settings. IFN-γ alone, one of the most frequently investigated markers, had the highest accuracy compared to the other cytokine signatures (AUC 0.89, 95% CI: 0.83–0.95; Figure 3d). However, this was lower than our combined signature, as well as our 3-cytokine signature (AUC 0.96, 95% CI: 0.92–0.99), although the confidence intervals overlapped (Figure 3d). The accuracies of the other cytokine signatures are shown in **Supplementary** Figure 3a. Unlike the metabolite signature, which showed improved performance in PLWH (Figure 3b), the IFN-γ cytokine signature performed consistently across HIV status (Figure 3e). As shown in **Supplementary** Figure 3c, TNF-α exhibited better performance in PLWH, whereas the accuracy of the IP-10 signature was reduced in this subgroup (**Supplementary** Figure 3b). Similar to the metabolite signature (i.e., Kynurenine/Tryptophan + Retinol), the IFN-γ cytokine signature demonstrated improved performance when hospitalized participants were excluded from the other respiratory disease control group (Figure 3f), and the same trend was observed for the other three cytokine signatures (**Supplementary Figures 3e, 3f** and 3g).

**Figure 3.**
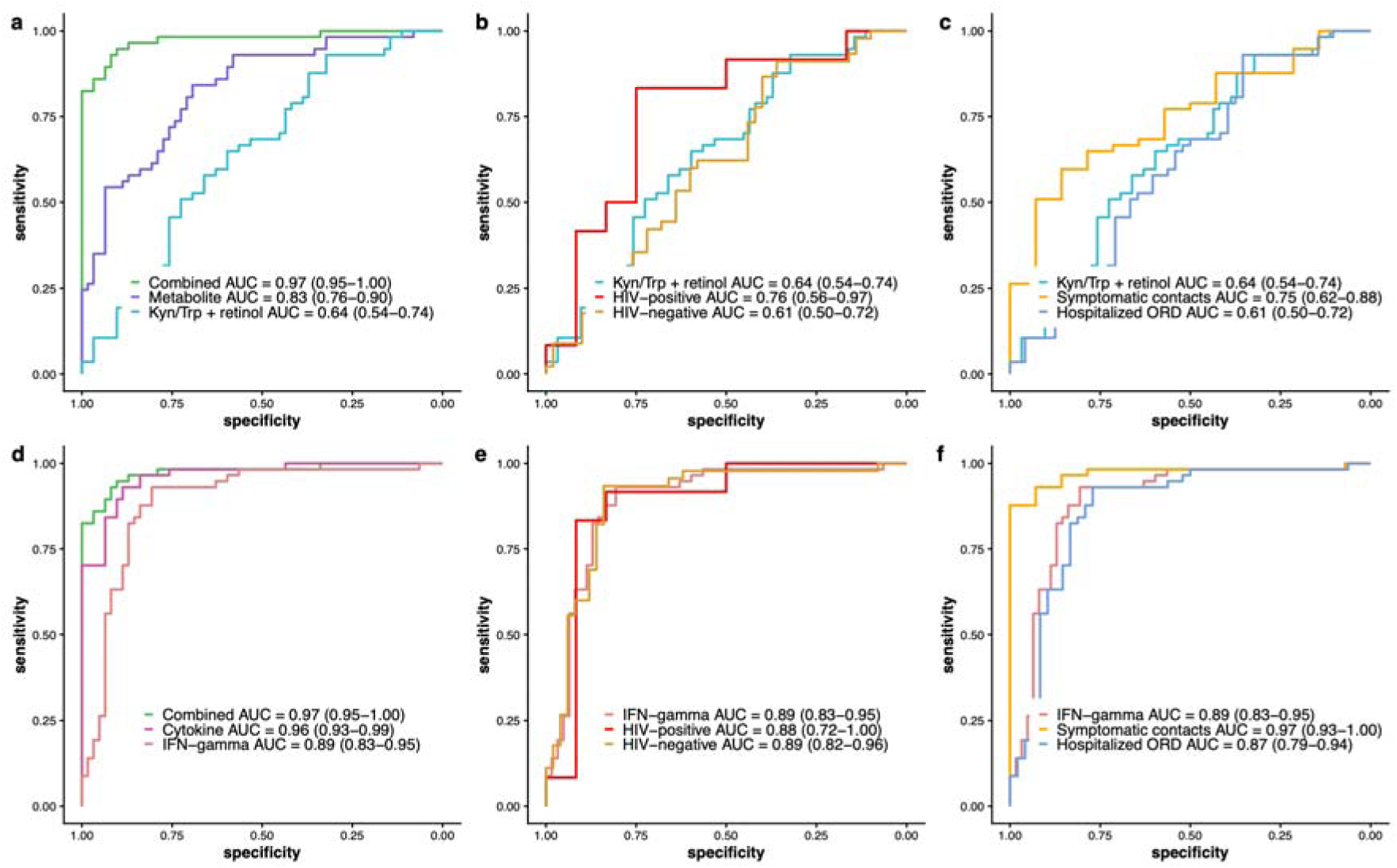
Comparison of our signature with published biomarker signatures for TB. (**6, 16, 18**)**. a)** ROC plots for the combined, metabolite, and Kyn/Trp + Ret signatures. **b, c)** Performance of the Kyn/Trp + Ret signature is further evaluated within subgroups defined by HIV status (HIV-positive and HIV-negative) and control type (symptomatic contacts and hospitalized LRTI) respectively. **d)** ROC plots for the combined, cytokine, and IFN-gamma signatures. **e, f)** Performance of the IFN-gamma is further evaluated within subgroups defined by HIV status (HIV-positive and HIV-negative) and control type (symptomatic contacts and hospitalized LRTI) respectively. AUC values with 95% confidence intervals are reported for all ROC analyses. ROC: Receiver Operating Characteristic; PTB: Pulmonary TB; AUC: Area Under the Curve; ORD: Other Respiratory Disease. Kyn: kynurenine; Trp: Tryptophan; Ret: Retinol.

### Identification of a multiomics signature for predicting PTB treatment failure

Out of 187 individuals with PTB, treatment outcomes were available for 178 participants, classified as either cured (n = 122) or failed treatment (n = 56). Out of 56 individuals who experienced treatment failure due to death (n = 17) or relapse/failure (n = 39), the majority were in the HIV-negative group (41/56). We repartitioned this PTB dataset into new discovery and validation datasets (Figure 1b), ensuring the same proportions of cured and failed cases as well as PLWH and HIV-negative individuals in both training and test sets. The new training dataset included 85 and 38 cases classified as cure and failure respectively (Figure 1b). This dataset was then used as input for *Boruta* (19) to identify potential biomarkers for predicting treatment outcome, which provided a more parsimonious feature selection list relative to *randomForestSRC*.

The Boruta algorithm selected six markers for predicting treatment outcome, including five metabolites (tyrosine, arachidonic acid, malate, eicosapentaenoic acid and valine) and one cytokine (IL.17A). We trained classification models using three sets of features: the combined 6-marker signature, the single-marker cytokine signature, and the 5-marker metabolite signature, with each model evaluated on the new test dataset. The combined and metabolite models had AUCs of 0.69 (95% CI: 0.54–0.84) and 0.70 (95% CI: 0.55–0.84), respectively, while the cytokine model performed worse (AUC = 0.56) (Figure 4a). Although none of the models demonstrated strong predictive performance, the results suggest that metabolites may have greater potential than cytokines in predicting treatment outcomes. Additionally, although a few metabolites and cytokines were differentially abundant between the cured and failed groups at the baseline, Week 8, or Week 17/26 (p-value < 0.05; **Supplementary** Figure 4, Figure 4b), none remained significant after multiple testing correction. This suggests that both metabolites and cytokines were not strongly associated with adverse treatment outcomes. Both the cytokine and metabolite models also showed poor performance in the HIV positive group, particularly at higher specificity thresholds (Figures 4d and 4e). However, only eight samples belonging to PLWH were included in this analysis, so validation in a larger dataset is needed.

**Figure 4.**
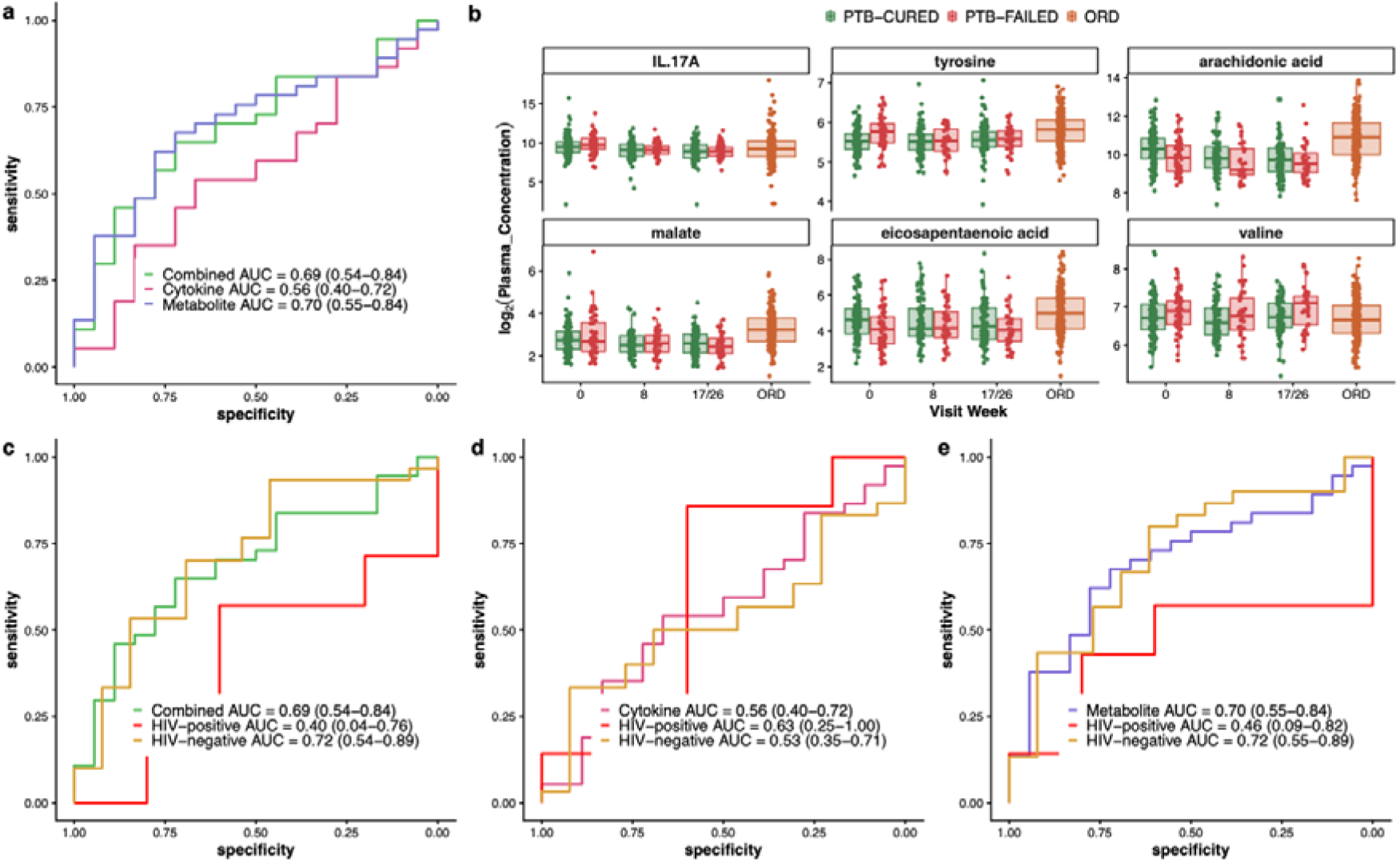
Identification of a multiomics signature for predicting TB treatment outcome. **a)** ROC curves for the combined, cytokine-only, and metabolite-only models. **b)** Longitudinal plasma concentrations of selected cytokines and metabolites identified by the *Boruta* algorithm at baseline (week 0), mid-treatment (week 8), and end of treatment (week 17 or 26). **c, d, e)** Performance of the combined, cytokine, and metabolite models within subgroups defined by HIV status (HIV-positive and HIV-negative). AUC values with 95% confidence intervals are reported for each ROC curve. ROC: Receiver Operating Characteristic; PTB: Pulmonary TB; ORD: Other Respiratory Disease; AUC: Area Under the Curve.

### Pathway enrichment

To identify metabolic pathways enriched in PTB versus other respiratory disease, we analyzed the full untargeted metabolomics data with both annotated and unannotated metabolites, which included 9,516 features in positive ion mode and 8,804 features in negative ion mode. We applied the linear model to each feature to calculate p-values for the PTB versus other respiratory disease comparison. As shown in Figure 5, in the Mummichog Python package, 25% of the top features in the null distribution model, 308 in positive mode (Figure 5a) and 567 in negative mode (Figure 2b), were considered significant (p-value < 0.0001) and used for pathway enrichment and module analysis. Out of 119 pathways, 22 unique pathways were significantly enriched (p-value < 0.05) in PTB versus other respiratory disease. Among these, 10 were in positive mode, 9 in negative mode, and 3 pathways were significantly enriched in both modes (Figure 5c). De novo fatty acid biosynthesis and fatty acid activation were the most significantly enriched pathways in PTB versus other respiratory disease (p-value < 0.0001), with nine out of ten and eleven metabolites in each pathway overlapping with our dataset, respectively. As illustrated in Figure 5e, the plasma concentrations of nine out of ten fatty acid metabolites included in the dataset were significantly lower in PTB compared to other respiratory disease (p-value < 0.0001), suggesting an increased utilization of fatty acids in PTB.

**Figure 5.**
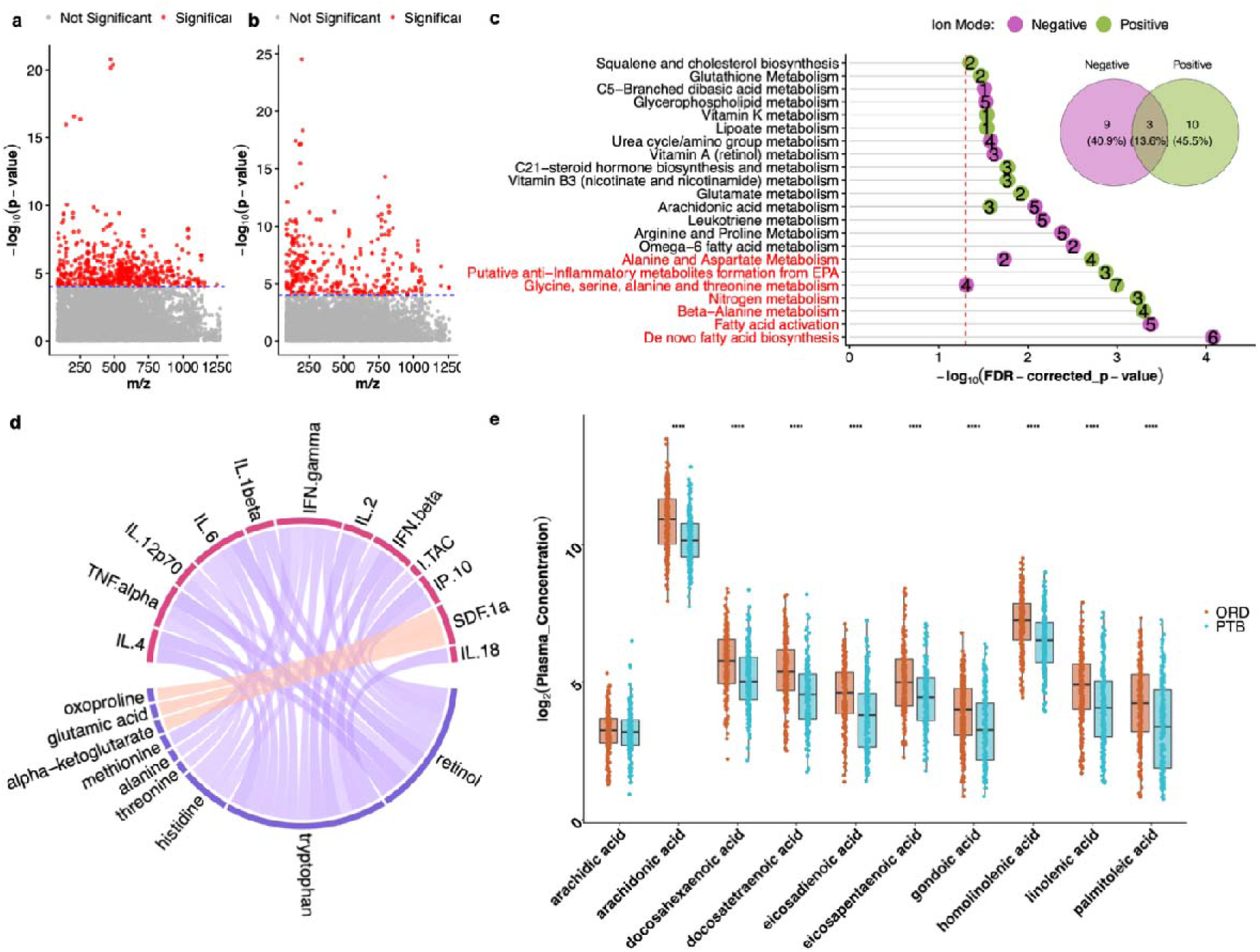
Summary of functional enrichment and integrative analysis. **a,b)** Manhattan plots showing significant metabolic features in positive and negative ion modes respectively, which were used as input for Mummichog to identify enriched pathways in the activity network. **c)** Bar plot of significantly enriched pathways in PTB versus ORD. Each row represents a pathway, with –log10(p-value) on the x-axis. The dashed red line indicates the significance threshold (p < 0.05), and the number within each circle indicates the overlap size between the pathway and the dataset. Green nodes correspond to positive ion mode results, and purple nodes to negative ion mode. A Venn diagram illustrates the overlap of enriched pathways identified in each ion mode. Pathways highlighted in red remained significant after FDR correction. **d)** Chord plot showing correlations between metabolites and cytokines. Only absolute correlations greater than 0.3 with p < 0.05 are shown. Red and blue chords indicate positive and negative correlations, respectively. **e)** Plasma concentrations of individual fatty acids compared between PTB and ORD groups. ORD: Other Respiratory Disease.

To determine associations between cytokine and metabolite signaling, we correlated plasma cytokine and metabolite concentrations across all samples. In the correlation analysis, we included only 22 cytokines and 46 metabolites that were differentially expressed (BH FDR-adjusted p < 0.05) between PTB and other respiratory disease (**Supplementary Figures 1** and 2). Among 1,012 correlation values, only 30 links showed significant correlations (BH FDR-adjusted p < 0.05 and absolute (Pearson correlation coefficient) > 0.3) and are presented in Figure 5d. All nine metabolites included in this plot are downregulated, while all cytokines except SDF-1a are upregulated in PTB compared to ORD (**Supplementary Figures 1** and 2). As illustrated in Figure 5d, all cytokines except SDF-1a are negatively correlated with the metabolites shown in the chord plot, whereas SDF-1a is positively correlated with alpha-ketoglutarate, glutamic acid, and oxoproline.

Tryptophan and retinol were significantly negatively correlated with 11 and 9 cytokines, respectively. Both metabolites are downregulated in PTB, and their reduced levels may be associated with the upregulation of multiple pro-inflammatory cytokines. Their broad negative correlations highlight their potential in modulating immune responses, with depletion contributing to enhanced inflammation in PTB. Among cytokines, IFN-γ, IL-6, TNF-alpha, and IFN-beta, each reported in multiple studies to be upregulated in PTB (20), are significantly negatively correlated with retinol, tryptophan, and histidine. This suggests that these four cytokines are particularly sensitive to metabolic perturbations and may act as central nodes linking metabolism and immune activation in PTB.

## Discussion

In this study, we identified a novel 5-marker signature comprising three cytokines and two metabolites for the diagnosis of PTB, which met the WHO TPP thresholds for both a non-sputum point-of-care (POC) TB diagnostic test and TB triage test. One of the key strengths of this signature is its consistent performance in subgroup analyses, including when the validation dataset was stratified by HIV status. Together, our findings highlight the innovation of a rapid, minimally invasive, plasma-based multiomics signature that has potential for translation into a POC platform. This would potentially enabling early, accurate, and accessible PTB diagnosis in diverse clinical settings.

Beyond the performance of the biomarker signature itself, several aspects of our study design and data analysis contribute to the robustness and reliability of our findings. First, our control group included both symptomatic contacts of index TB cases and hospitalized participants with other non-TB respiratory illnesses. Many studies have used asymptomatic or minimally symptomatic participants in control groups, potentially overestimating ROC performance by identifying general markers of inflammation that are not specific to TB (21–23). Our evaluation of previously published signatures showed reduced performance when hospitalized participants were included in the control group. Conversely, our signature performed well when the control group in the validation dataset included only symptomatic contacts or only hospitalized participants. Second, unlike most studies that analyzed cytokines or metabolites separately, we integrated both data types, which improved the accuracy of the signature at higher specificity thresholds. Finally, although many biomarker discovery studies report ROC results only in the training cohort, which often leads to overestimation of AUC results, the sample size in our study was sufficient to divide participants into independent training and validation cohorts. This allowed us to evaluate signature performance exclusively in the validation set.

Next, we also examined which component of the signature contributed most to its performance. We observed that most of the signal in the combined signature was driven by cytokines. All three cytokines included in our 5-marker signature were previously identified as biomarkers for PTB, either individually or in combination with other cytokines (6). While IFN.gamma and IL.22 were upregulated in PTB compared with other respiratory disease, IL.10 showed downregulation in our study, whereas it is often reported as upregulated in TB. We found that this discrepancy is due to the inclusion of hospitalized pneumonia patients in the control group, who exhibited higher IL.10 levels compared with PTB cases. This illustrates the importance of using control groups with a variety of symptom severity. While IFN.gamma and IL.10 have been extensively investigated in hypothesis-driven TB biomarker studies(6), IL.22 has only been reported in one study, in combination with sCD40L, for diagnosing TB pleurisy (24).

In addition to the cytokines in our 5-marker signature, the inclusion of two metabolites, methionine and oxoproline, further enhanced the signature’s performance, particularly at higher specificity thresholds. This improvement may arise because metabolite measurements capture additional TB-specific signaling pathways beyond the acute inflammatory response reflected by cytokines. Consistent with our findings, methionine, together with three other metabolites, demonstrated good performance in distinguishing TB from latent infection and healthy controls using plasma samples (25). Another targeted metabolomics study also identified methionine as a metabolite decreased in the serum of TB patients compared with individuals with latent TB infection and healthy controls (16). Like methionine, oxoproline was also observed at decreased levels in the serum of TB patients compared with healthy controls and has been suggested as a potential TB biomarker (26).

In addition to identifying biomarkers for TB diagnosis, we also investigated whether cytokines and metabolites could predict TB treatment failure. In contrast to their strong performance in TB diagnosis, cytokines showed poor predictive ability for treatment outcomes, whereas metabolites demonstrated better performance. Among all metabolites, a high concentration of many unsaturated fatty acids was associated with treatment success, which suggests their potential clinical value as biomarkers for predicting treatment response and guiding patient stratification and monitoring.

To our knowledge, few studies measured plasma metabolomics at multiple timepoints during TB treatment (17). However, our findings are supported by previous studies, which underscore the prognostic value of baseline metabolites. For example, one study (27) analyzed urine samples collected at baseline (pre-treatment) from TB patients with successful versus unsuccessful treatment outcomes and identified metabolites predictive of failure, having an AUC of approximately 0.94.

We also observed that many long chain fatty acids were significantly decreased in PTB compared to other respiratory disease and pathways related to fatty acid metabolism were significantly enriched in PTB versus other respiratory disease. This enrichment highlights a key feature of the host metabolic response to Mtb infection and disease, shared between humans and non-human primates, characterized by a systemic depletion of host lipids (28). However, the mechanism for this depletion remains unknown. Infected macrophages often accumulate lipid droplets, providing nutrients for the bacteria (29), which may contribute to increased utilization of host lipids. Supporting this, previous studies have demonstrated that macrophages unable to import, store, or catabolize fatty acids restrict *M. tuberculosis* growth (30). By contrast, lipid metabolic alterations have also been shown to regulate lung inflammation in other respiratory infections (31).

Additionally, we found significant negative correlations between the metabolites tryptophan, retinol, and histidine with multiple proinflammatory cytokines, and all three metabolites were downregulated in PTB consistent with other studies (32–34). The tryptophan catabolism pathway, driven by upregulation of the enzyme indoleamine 2,3-dioxygenase-1 (IDO-1), has been shown to reflect disease activity in TB, with an elevated kynurenine/tryptophan ratio correlating with immune activation and cytokine signaling (17). In addition, studies of retinol metabolism demonstrate that its conversion to all-trans-retinoic acid (ATRA) in dendritic cells can stimulate macrophage antimicrobial responses and contribute to immune activation during TB infection (35). The downregulation of histidine observed in our PTB cohort is consistent with previous reports showing reduced plasma histidine levels at the time of TB diagnosis and normalization following treatment. This may reflect host-mediated histidine depletion via IFN-γ-induced catabolic pathways, which are thought to restrict pathogen access to this essential amino acid (36).

Our study was subject to limitations. First, nearly all participants in our study were enrolled after presenting for clinical care with respiratory symptoms, so it is unknown how the signature may perform in community screening settings where sub-clinical TB is more common. Second, we did not investigate the performance of the signature for diagnosing extrapulmonary TB. Third, although participants were enrolled at multiple sites, most participants were from South Africa, resulting in a limited geographical distribution. Given the heterogeneity of TB across different regions, the generalizability of the signature needs to be validated in additional populations.

In summary, our study demonstrates that a combined cytokine-metabolite signature can reliably distinguish PTB from other respiratory disease, meeting WHO TPP thresholds for both diagnostic and triage applications. While cytokines largely drive the inflammatory signal, metabolites enhance sensitivity, particularly at higher specificity thresholds, and provide additional prognostic value for treatment outcomes. Our findings support the potential of integrated multiomics approaches for both early TB diagnosis and monitoring treatment response, particularly in clinically relevant settings such as symptomatic or hospitalized populations. These insights lay a foundation for translating multiomics biomarker signatures to POC platforms for TB diagnosis.

## Methods

### Ethical considerations

Written informed consent was obtained from all study participants, and study approval was obtained from the institutional review boards (IRBs) of Emory University (Atlanta, Georgia, USA, IRB # 00006763) and the Human Research Ethics Committee (Medical) of the University of the Witwatersrand(Wits HREC:171009), South Africa.

### Cohort

The plasma samples used in this study were collected from three distinct biorepositories. A subset of participants with TB disease (n=119) were analysed from the MARK-TB biorepository. This biorepository included samples from participants from two randomized controlled trials of treatment-shortening regimens for drug susceptible TB disease and participants enrolled in a longitudinal AIDS Clinical Trial Group Study (12, 13). Full protocols for the parent studies were published previously. All samples analysed were from participants at least 18 years of age diagnosed with PTB by a positive sputum culture for Mtb or Xpert MTB result.

Additional plasma samples were obtained from 233 adult patients hospitalized with a respiratory illness within 48 hours of admission in South Africa (14). Of these, 68 had microbiologically confirmed PTB. Of the remaining 165 in whom PTB was excluded, most were diagnosed with pneumonia (n=126), while others were diagnosed with an acute pulmonary embolism or exacerbations of asthma, chronic obstructive pulmonary disease, or heart failure (14).

Finally, outpatient controls were enrolled from a South African study of close household contacts of persons with confirmed TB disease (n=39). We included participants who were at least 18 years of age reporting at least one TB symptoms (cough, fever, weight loss or night sweats) at the time of evaluation with negative sputum Mtb culture and Xpert MTB results (37).

Out of 187 individuals with PTB, treatment outcomes were available for 178 participants, classified as either cured (n = 122) or failed treatment (n = 56). All individuals who experienced treatment failure, relapse, or death due to TB were classified as failures. Some PTB individuals were followed longitudinally during treatment, with plasma samples available at mid-treatment (week 8, n=116) and at the end of treatment (week 17/26, n=119).

### Metabolomics analysis

High-resolution plasma metabolomics was performed using a standardized method, which has been published previously (17, 38, 39). Briefly, 50 μL of thawed plasma was treated with 100 μl acetonitrile (2:1, v/v) and an internal isotopic standard mixture (3.5 μL/sample) consisting of 14 stable isotopic chemicals for quality control (40). Samples were analyzed in triplicate using an Orbitrap Q Exactive Mass Spectrometer (Thermo Scientific, San Jose, CA, USA) with dual liquid chromatography methods: Hydrophilic Interaction Liquid Chromatography (HILIC) and C18 reversed phase chromatography (Higgins Analytical, Targa, Mountain View, CA, USA, 2.1 x 10 cm) with a formic acid/acetonitrile gradient followed by positive or negative electrospray ionization, respectively The mass spectrometer was operated over a scan range of 85 to 1275 mass/charge (*m/z*) and data was stored as .Raw files (41). Data were extracted and aligned using apLCMS (42) and xMSanalyzer (43) with each feature defined by specific *m/z* value, retention time, and integrated ion intensity (41). Identities of targeted metabolites were confirmed and quantified by accurate mass, and retention time relative to an in-house library of authentic standards (44).

### Plasma cytokine detection

The U-PLEX assay (Meso Scale MULTI-ARRAY Technology) commercially available by Meso Scale Discovery (MSD) was used for plasma cytokine detection. This technology allows the evaluation of multiplexed biomarkers by using custom made S-PLEX and U-PLEX sandwich antibodies with a SULFO-TAG™ conjugated antibody and next generation of electrochemiluminescence (ECL) detection. The assay was performed according to the manufacturer’s instructions. In summary, 25μL of plasma from each participant was combined with the biotinylated antibody plus the assigned linker and the SULFO-TAG™ conjugated detection antibody; in parallel a multi-analyte calibrator standard was prepared by doing 4-fold serial dilutions. Both samples and calibrators were mixed with the Read buffer and loaded in a 10-spot U-PLEX plate, which was read by the MESO QuickPlex SQ 120. The plasma cytokines values (pg/mL) were extrapolated from the standard curve of each specific analyte.

### Metabolite and cytokine abundant data analysis

We applied the *lmFit* and *eBayes* functions from the *limma* R package (45) to estimate log2 fold changes and corresponding p-values (adjusted for multiple testing using the Benjamini–Hochberg false discovery rate (FDR) method (46)) for metabolite and cytokine plasma concentration levels between PTB and other respiratory disease groups. Age, sex, and HIV status were included as covariates in the differential analysis of metabolites and cytokines. To illustrate variation across samples within each dataset, we performed Principal Component Analysis (PCA) using the prcomp function in R. Heatmaps were generated using the *ComplexHeatmap* R package(47), and other plots were created with the *ggplot2* R package (48). Additionally, correlations between the metabolite dataset and cytokine dataset were visualized using a chord diagram, with only correlations showing p-values < 0.05 and absolute Pearson correlation coefficient > 0.3 included in the plot.

### Multiomics biomarker discovery analysis

To identify multiomics biomarkers for TB diagnosis, we applied *randomforestSRC* (15), a feature selection approach based on an ensemble of decision trees, to the combined metabolite and cytokine dataset across all samples. The performance of each signature was evaluated using a support vector machine (SVM) (49) with a radial basis function (RBF) kernel, implemented via the *svmRadial* method in the *caret* R package. This approach is well-suited for capturing the heterogeneous and potentially non-linear relationships typical of multiomics data. Model performance was assessed by calculating the area under the receiver operating characteristic curve (AUC-ROC) with corresponding 95% confidence intervals (CIs). ROC curves were used to illustrate the sensitivity and specificity of each signature in distinguishing PTB from other respiratory disease across a range of thresholds. To compare the multiomics models against single-omics models, we also trained SVM classifier separately using only metabolite and cytokine features. We also used the *Boruta* approach (19), another feature-selection method based on random forests, to identify the metabolites and cytokines with the greatest predictive power for treatment-outcome prediction.

### Untargeted metabolomics pathway enrichment analysis

In the untargeted LC-MS/MS feature dataset, p-values for each feature were calculated using a linear model adjusted for sex, age, and HIV status to compare PTB and other respiratory disease samples. Features, along with their *m/z* values, retention times, and p-values, were then provided as input to Mummichog (50), a Python package for pathway and network enrichment analysis in metabolomics that does not require prior metabolite identification. Mummichog maps significant features (the top 25% of features with p-values < 0.05 in the null distribution model) to all possible candidate metabolites, and projects them onto known metabolic networks from the KEGG database (51). It then evaluates whether clusters of mapped metabolites are enriched in specific pathways compared to random expectations. Features mapping to significantly enriched clusters are considered more likely to represent biologically relevant metabolites.

## Code Availability

All code necessary for the data analysis and visualization is available at https://github.com/zmousavian/Multiomics-TB-biomarker. Any additional information required to reanalyze the data reported in this paper is available from the lead contact upon request.

## Data Availability

Data is provided within the supplementary information files.

## Acknowledgements

The authors are grateful to all study participants, and to the clinicians and staff at our multiple clinical sites who provided care for the participants in this study. This work was supported by grants from the U.S. National Institute of Allergy and Infectious Diseases (NIAID) [R01 AI182244, R21 AI178324, K23 AI144040, P30 AI168386, P30 AI050409, K24 AI114444]; and the National Center for Advancing Translational Sciences [UL1 TR002378], Bethesda, MD, USA.

## Author Contributions

Conceptualization: ZM, NRG, MJM, NM, JMC; Data curation: ZM, FN, MN, MI, MJM, JMC; Formal analysis: ZM, MJM, JMC; Funding acquisition: RRK, MJM, JMC; Investigation: All authors; Methodology: ZM, MJM, DPJ, AAS, JMC; Project administration: ZM, MJM, JMC; Software: ZM; Resources: ZM, MJM, DPJ, AAS, JMC; Supervision: ZM, MJM, NM, AAS, JMC; Validation: ZM, MJM, JMC; Visualization: ZM; Writing–original draft: ZM, MJM, JMC; Writing–review & editing: All authors.

## Competing interests

The authors declare no competing interests.

## Notes

### Competing Interest Statement

The authors have declared no competing interest.

