## Supplementary Materials for "Plasma multiomics distinguishes pulmonary tuberculosis from other respiratory infections"

**Jeffrey M. Collins**

J

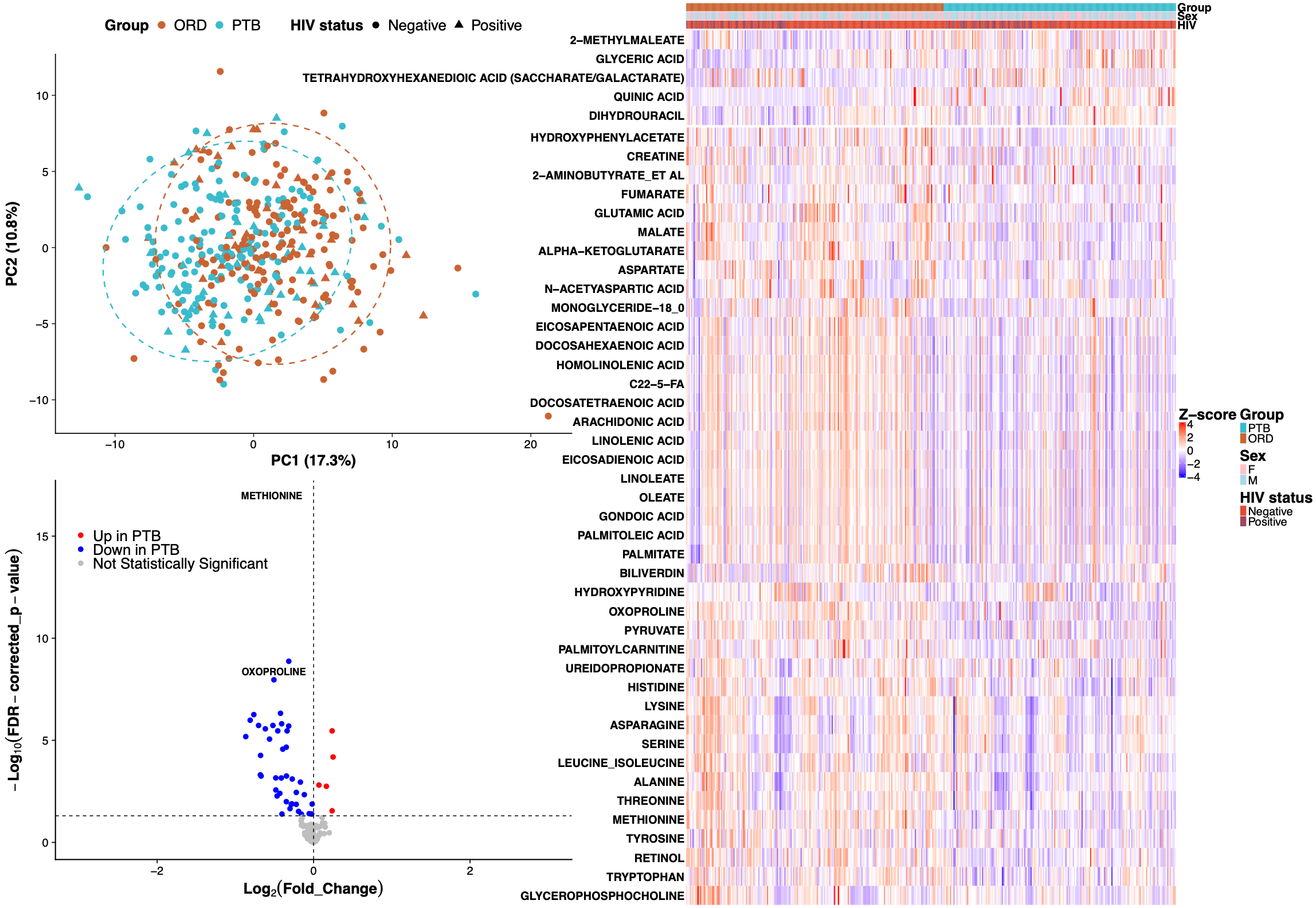

**Supplementary Figure 1.** Principal Component Analysis (PCA), volcano plot, and heatmap of the metabolite dataset. The PCA plot illustrates overall variation and sample clustering based on metabolite profiles. The volcano plot highlights differentially abundant metabolites between pulmonary TB (PTB) and other respiratory diseases (ORD), with significance defined as FDR-adjusted p-value < 0.05. Red points indicate metabolites upregulated in PTB, while blue points indicate those downregulated in PTB. The heatmap shows the relative abundance of all metabolites that were significantly differentially regulated between the PTB and ORD groups.

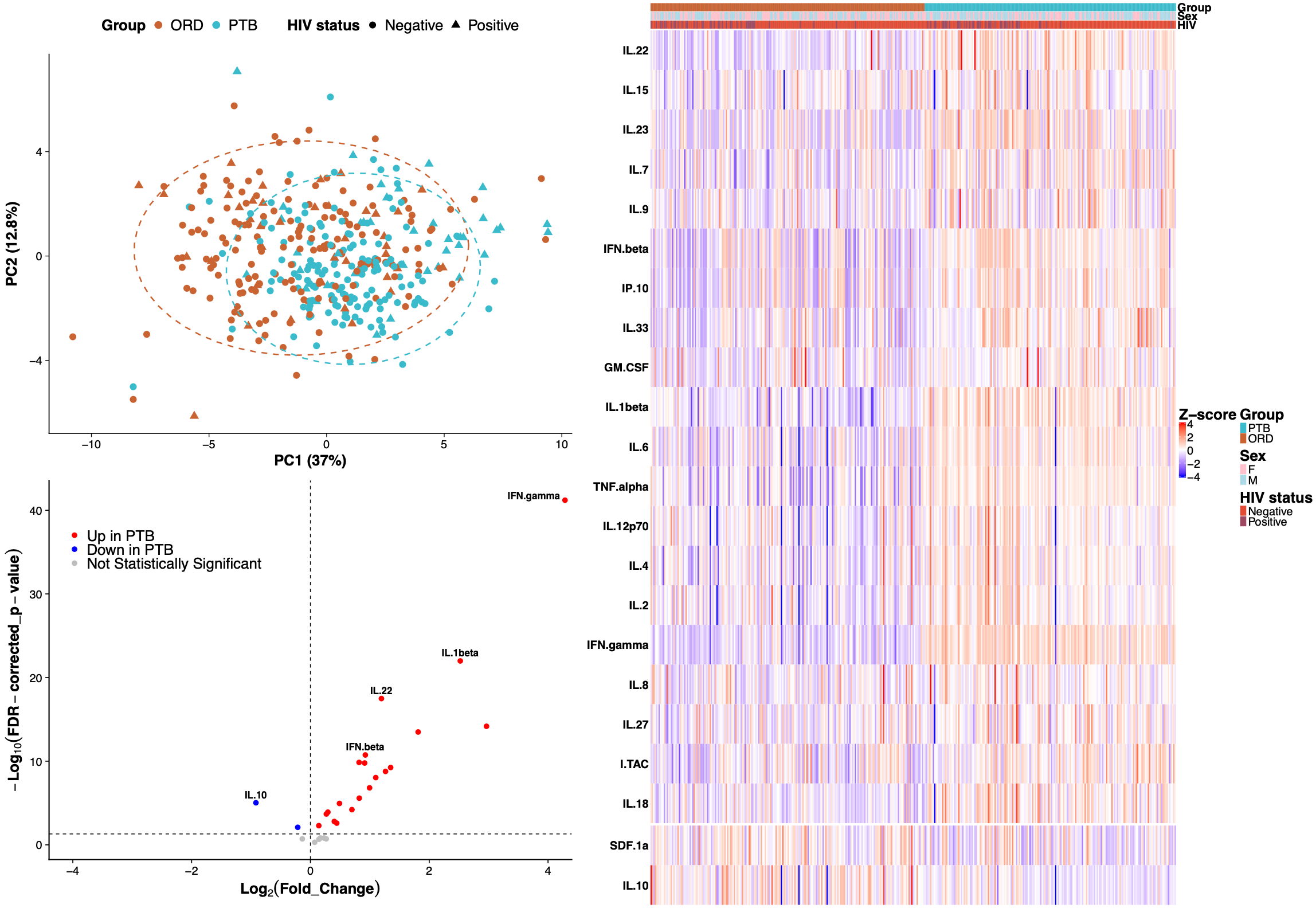

**Supplementary Figure 2.** Principal Component Analysis (PCA), volcano plot, and heatmap of the cytokine dataset. The PCA plot illustrates overall variation and sample clustering based on cytokine profiles. The volcano plot highlights differentially abundant cytokines between pulmonary TB (PTB) and other respiratory diseases (ORD), with significance defined as FDR-adjusted p-value < 0.05. Red points indicate cytokines upregulated in PTB, while blue points indicate those downregulated in PTB. The heatmap shows the relative abundance of all cytokines that were significantly differentially regulated between the PTB and ORD groups.

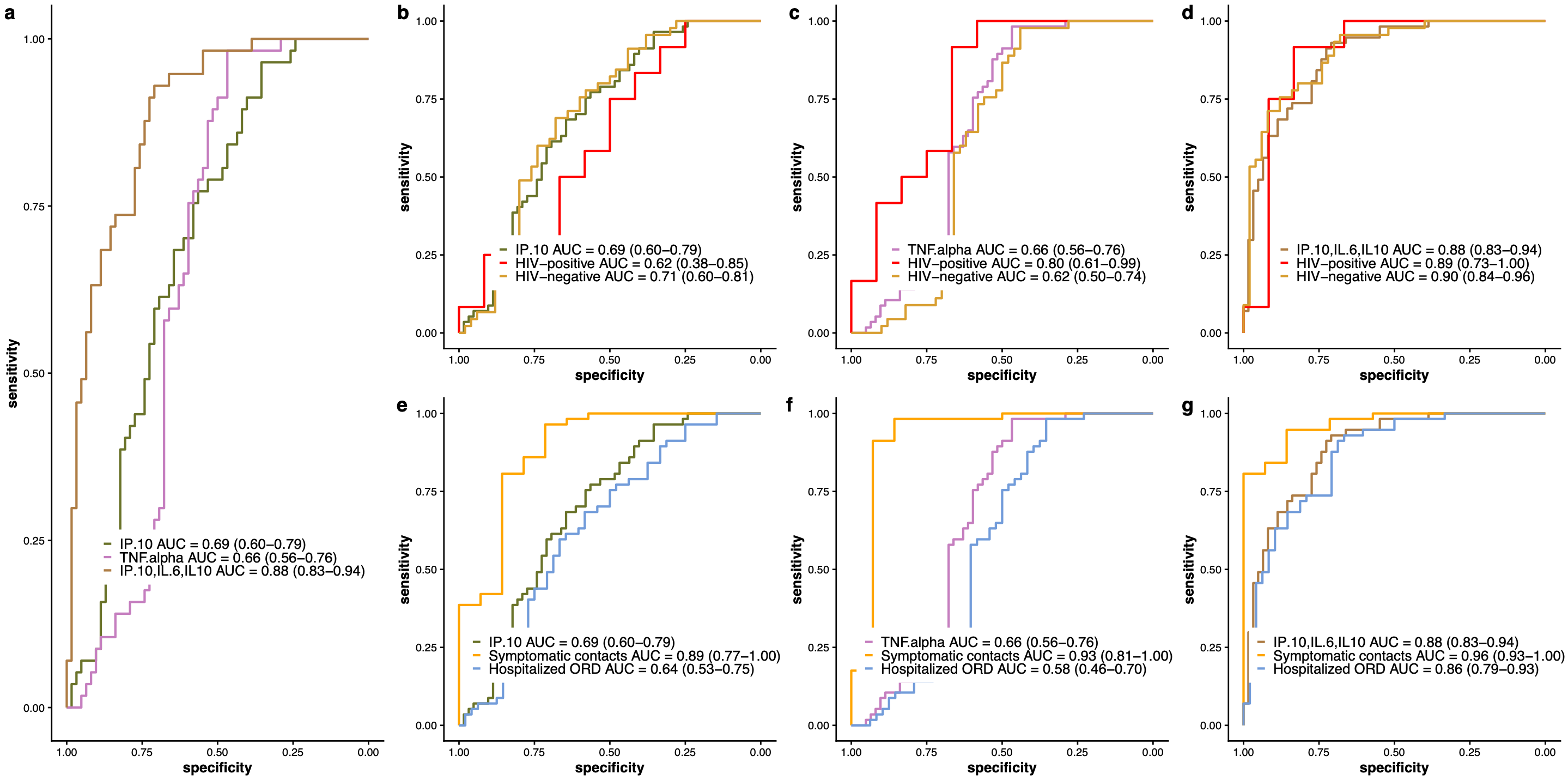

**Supplementary Figure 3.** Receiver Operating Characteristic (ROC) curves for three additional cytokine signatures: (1) IP-10, (2) TNF-α, and (3) a 3-marker signature consisting of IP-10, IL-6, and IL-10. These signatures were evaluated for their ability to distinguish Pulmonary TB from other respiratory disease. ROC analyses are presented for the overall dataset (**a**), as well as for subgroups stratified by HIV status (HIV-positive and HIV-negative) (**b, c, d**) and control type (symptomatic contacts and hospitalized LRTI cases) (**e, f, g**). Area under the curve (AUC) values with 95% confidence intervals are reported for each model and subgroup. ORD: Other Respiratory Disease; AUC: Area Under the Curve.

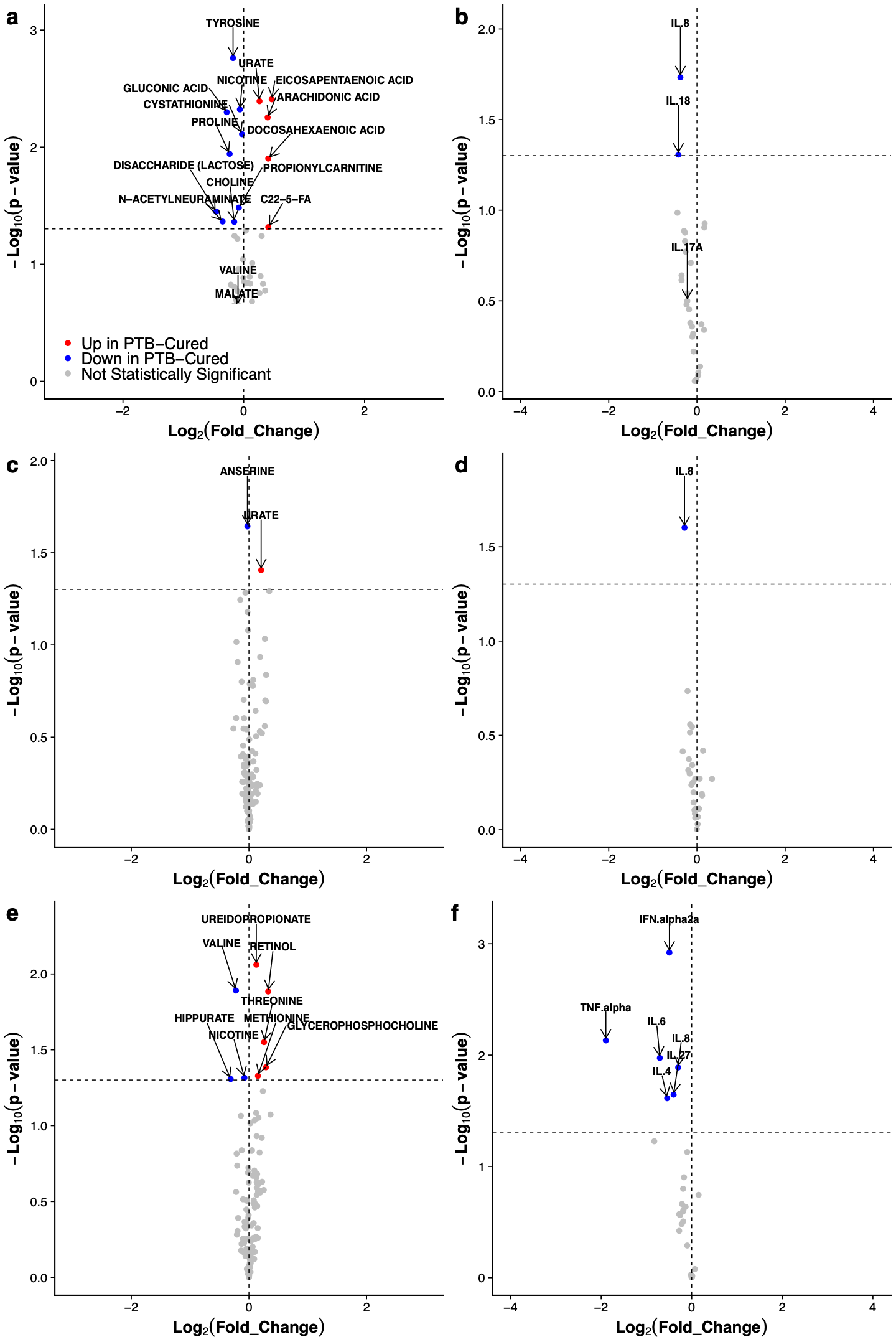

**Supplementary Figure 4.** **a, c, e)** **Volcano plots highlight differentially abundant metabolites in cure versus failure groups at baseline, mid-treatment (week 8), and end of treatment (week 17 or 26) respectively, with significance defined as p-value < 0.05. b, d, f)** **Volcano plots highlight differentially abundant cytokines in cure versus failure groups at baseline, mid-treatment (week 8), and end of treatment (week 17 or 26) respectively, with significance defined as p-value < 0.05. Red points indicate markers upregulated in the cure group, blue points indicate those downregulated in the cure group, and gray points represent non-significant markers.**
